# The minimum number of blood pressure measurements needed and thresholds for visit-to-visit blood pressure variability to predict cardiovascular disease in primary care patients

**DOI:** 10.64898/2026.03.02.26347458

**Authors:** Mifetika Lukitasari, Reza Argha, Siaw-Teng Liaw, Bin Jalaludin, Joel Rhee, Jitendra Jonnagaddala

**Affiliations:** School of Population Health, UNSW Sydney, Australia; School of Nursing, Faculty of Health Science, Brawijaya University, Indonesia; Graduate School of Biomedical Engineering, UNSW Sydney, Australia; Discipline of General Practice, School of Clinical Medicine, UNSW Sydney, Australia; SREDH Consortium, UNSW Sydney, Australia

**Keywords:** Visit-to-visit Blood Pressure Variability, Cut-off, Electronic Health Records, Cardiovascular Disease, Risk Assessment

## Abstract

**Objectives:** Visit-to-visit blood pressure variability (VVV BPV) is an underutilised risk factor for cardiovascular disease (CVD). This study aims to determine the minimum number of BP measurements needed and to identify cut-off values for the standard deviation (SD), coefficient of variation (CV), and average real variability (ARV) of systolic and diastolic VVV BPV to predict CVD risk in primary care.

**Methods:** We analysed data from the electronic practice-based research network (ePBRN) in Southwestern Sydney, including patients aged 18-55 with at least eight BP readings. Patients with incomplete data or no follow-up beyond age 55 were excluded. The agreement between SD calculated from 3-5 measurements and 8 measurements (reference) was evaluated using Pearson’s correlation coefficient and the intraclass correlation coefficient. Then, after identifying that a minimum of five BP measurements is needed, another cohort with at least five BP measurements was developed. Percentile-based cut-offs (10^th^ – 90 ^th^, 5-percentile increments) were derived for systolic and diastolic BPV (SD, CV, ARV). Predictive accuracy was assessed using the C-statistic. The outcome was the first CVD occurrence.

**Results:** A total of 1,549 patients were included in the first study. Five BP measurements showed good agreement with eight measurements (ICC: 0.79; correlation: 0.80). A total of 3,022 patients were included (55.2% women). Higher VVV BPV (SD, CV. ARV) was associated with increased CVD risk. Optimal cut-off values for systolic BP were 19 mmHg (SD), 14% (CV), and 15 mmHg (ARV), and for diastolic BP were 11 mmHg (SD), 12% (CV), and 11 mmHg (ARV). Predictive performance was consistent across time frames.

**Conclusions:** These BPV cut-offs provide clinically relevant thresholds for CVD risk prediction. At least five BP measurements are sufficient to estimate BPV for this purpose.

## Introduction

Elevated blood pressure (BP) is an important risk factor for cardiovascular disease (CVD) morbidity and mortality.^1^ However, BP readings recorded during clinic visits can vary considerably and fluctuate from appointment to appointment due to various factors. Recently, interest has shifted from merely categorising individual BP measurements to analysing BP variability across clinical visits. This variability, termed visit-to-visit blood pressure variability (VVV BPV), is an emerging crucial risk factor for CVD.^2^

BP variability is generally viewed as a normal physiological phenomenon.^3^ However, an increasing number of studies, including clinical trials, observational studies, and analyses of electronic health records (EHR), have shown that increased VVV BPV is linked to increased cardiovascular events and mortality.^4^ Previous meta-analyses have shown that this variability can serve as a predictor of cardiovascular disease and death, irrespective of comorbidities or patient demographics.^5^

Previous research on visit-to-visit BPV in Australia has primarily drawn on data obtained through the Second Australian National Blood Pressure Study (ANBP2) ^6^ and the ASPirin in Reducing Events in the Elderly (ASPREE) study.^7^ Both studies are anchored in clinical trial data, which provides a controlled environment for understanding BPV. However, in real-world clinical practice, BP data from office measurements guide the primary prevention and management of CVD. EHR data, characterised by their real-world applicability, offer a comprehensive, longitudinal perspective on patients’ health profiles, capturing various factors such as comorbidities, treatment regimens, and lifestyle choices that may influence BP.

CVD remains a leading cause of death in Australia.^8^ BP data from EHR can be used to predict CVD by calculating VVV BPV. This approach offers a valuable method for improving clinical practices and public health strategies. While previous meta-analyses provided valuable insights into the association between VVV BPV and CVD.^4,5,9,10^ A significant gap persists in identifying a definitive cutoff for BPV from office BP readings that reliably predict CVD risk outside clinical trials. This absence of evidence underscores the need for further research to define the VVV BPV thresholds as predictive markers of CVD across diverse demographic groups. Additionally, to ensure the VVV BPV cut-off accuracy, it is essential to determine the minimum number of BP measurements needed for a reliable estimate.

To our knowledge, this is the first study to establish cut-off values for VVV BPV in predicting CVD. Initially, it aims to identify the minimum number of BP measurements needed to reliably calculate VVV BPV for predicting CVD events. Subsequently, based on the findings from this first step, the study seeks to develop optimal cut-off values for the standard deviation (SD), coefficient of variation (CV), and average real variability (ARV) of VVV BPV as predictors of CVD, following an investigation into the relationship between VVV BPV and the occurrence of initial CVD events in the general population.

## Methods

This study was conducted in accordance with the principles outlined in the Transparent Reporting of a multivariable prediction model for Individual Prognosis or Diagnosis (TRIPOD)^11^ as presented in Table S1. It received approval from the Human Research Ethics Committee of the University of New South Wales, Sydney, Australia (approval number: HC230072). This study comprises two main parts: the first to determine the minimum number of BP measurements needed for VVV BPV calculation, and the second to identify the cut-off value of VVV BPV for predicting CVD.

### Study Design

We used the electronic practice-based research network (ePBRN) dataset, derived from a network of consented General Practitioner (GP) clinics and secondary/tertiary hospitals in Southwestern Sydney, Australia. The data were entered into EHR systems by clinicians and care providers, including GPs, hospital doctors, allied health professionals, and nurses, during patient care from 2006 to 2019. This routinely collected data was subsequently extracted, de-identified, and linked using a third-party tool (GRHANITE™ linkage).

### Participants

We used a landmark point (study baseline) at age 55 years in both studies; the study timeline is presented in Figure S1. Therefore, the inclusion criteria were as follows: 1) Patients aged 18 to 55 years with recorded BP measurements; 2) No prior diagnosis of atherosclerotic CVD or atrial fibrillation at the index date, defined as the patient’s 55^th^ birthday. 3) At least eight BP readings obtained during separate visits for the first study, with a minimum of two weeks between each visit; Exclusion criteria included patients with no recorded sex or year of birth, as well as those without any visits after age 55.

Of the 20,757 patients with linked general practice and hospital data, we excluded 7,870 patients who had fewer than eight BP readings before age 55. We also excluded 10,987 patients without outcome data available after age 55, and 351 patients with outcome data recorded before age 55. This left 1,549 patients for analysis of the first study. Later, after completing the analysis of the first study, we determined that at least five BP measurements are needed to calculate VVV BPV for predicting CVD. We further filtered the cohort for the second study to include only those with at least five BP measurements, resulting in 3,022 patients eligible for analysis. The flow chart of cohort selection is presented in Figure S2.

### Power calculation

Using the established formula for calculating the sample size in Cox regression analysis, we estimated the required sample size based on an expected event rate of 6.7% ^8^. For a one-SD increase in each BPV metric, the study achieved a statistical power of 0.90 with a sample size of 3,022 and a two-sided alpha value of 0.05.

### Outcome

The outcome of interest was the first CVD event diagnosis within the cohort. A CVD event was defined as fatal and non-fatal coronary heart disease (including myocardial infarction and angina), stroke or transient ischemic attack, heart failure, atrial fibrillation, or peripheral arterial disease. The incidence of CVD was determined from the first CVD event in either dataset (GP or hospital).

### Predictors

The following BP measurements were considered biologically implausible and thus excluded: SBP <60 mmHg, SBP >250 mmHg, DBP <40 mmHg, and DBP > 140 mmHg. We used the SD of VVV BPV as the primary predictor of CVD in the main analysis. Additional analyses were conducted using the CV and ARV as alternative VVV BPV measures.

### Statistical analysis

#### Determining the minimum number of blood pressure measurements to calculate the BPV for predicting CVD

VVV BPV was calculated using the SD of the first 3 to 8 BP measurements. The agreement between the SDs of each subset and the SD of 8 measurements (used as the reference) was evaluated using Pearson’s correlation coefficient and the intraclass correlation coefficient (ICC). Eight measurements were used as a reference due to methodological feasibility; beyond that, the number of included patients in this study dropped below 1,000. Higher correlation coefficients indicate greater agreement. ICC values are interpreted as follows: <0.5 indicates poor reliability, 0.5-0.75 moderate reliability, 0.75-0.90 good reliability, and >0.90 excellent reliability.^12^

### Model development

The Cox proportional hazards model was used to estimate hazard ratios (HRs) for CVD development. The time-to-event was defined as the time between a patient’s 55^th^ birthday and either the date of the first CVD diagnosis or the censoring date. Descriptive statistics are presented as means with SDs and proportions, as appropriate. The main analysis estimated HRs based on a one-SD increase in each BPV metric. Participants without CVD events were censored at their last recorded contact on or before December 2019. Five models were developed: Model 1: unadjusted; Model 2: adjusted for sex and mean BP; Model 3: adjusted for sex, mean BP, and comorbidities; Model 4: adjusted for sex, mean BP, comorbidities, and laboratory values, including total cholesterol/HDL ratio, estimated glomerular filtration rate (eGFR), fasting blood glucose, and body mass index (BMI); Model 5: adjusted for sex, mean BP, comorbidities, laboratory values (total cholesterol/HDL ratio, eGFR, fasting blood glucose, and BMI), and medications (antihypertensive drugs, lipid-lowering agents, anticoagulants, and anti-diabetes medications as presented in Table S2). The proportional hazards assumption was assessed using the Schoenfeld residual plot. The Benjamini-Hochberg (BH) adjustment was used to control the false discovery rate in multiple statistical tests (Model 5).

We performed multiple imputations using chained equations (MICE)^13^ To address missing predictor data, we generated 20 imputed datasets. The association was analysed using Cox proportional hazard from each imputed dataset, and the pooled HR was extracted. All imputations and analyses were conducted in R version 4.4.0, using the MICE and survival packages (version 3.7-0) for Cox regression modelling.

To establish percentile-based cut-off values for systolic and diastolic BPV for each BPV metric (SD, CV, and ARV), we calculated percentiles from the 10th to the 90th, in 5-percentile increments (i.e., 10th, 15th, 20th, …, 90th). These cut-off values were subsequently used to predict outcomes.

The predictive performance of each cut-off was evaluated using the concordance index (C-statistic), a measure of model discrimination analogous to the area under the curve (AUC). C-statistic values ranged from 0.5, indicating no discriminative ability, to 1, indicating perfect discrimination. The final cut-off value for each BPV metric was determined based on the highest observed C-statistic. Additionally, the time-varying AUC was calculated to assess predictive performance for CVD over 2-, 4-, and 5-year time horizons.

## Results

In the first study, the correlation of the SD of systolic BP measurements increased progressively with the number of BP readings, as shown in Figure S3. Comparing 3, 4, 5, 6 and 7 systolic BP readings with eight systolic BP readings, the correlation coefficient progressively increased from 0.47 to 0.97 while the ICC increased from 0.46 to 0.97. Five systolic BP measurements showed good agreement with eight systolic BP measurements, as indicated by an ICC of 0.79 and a correlation of 0.80.

The second study included patients aged 18-55 years (55.2% women), with baseline characteristics presented in Table 1. The duration of BP observation was 5.74 ± 3.82 years. Before the age of 55, mean ± SD systolic and diastolic BP (mmHg) were 136.0 ± 16.0 and 82.0 ± 7.9, respectively. Approximately, 34.8% were smokers, 6.8% had diabetes mellitus, 32.8% had hypertension, and 18.4% had dyslipidaemia. Over a mean follow-up of 5.2 years, 231 patients developed CVD.

**Table 1.**
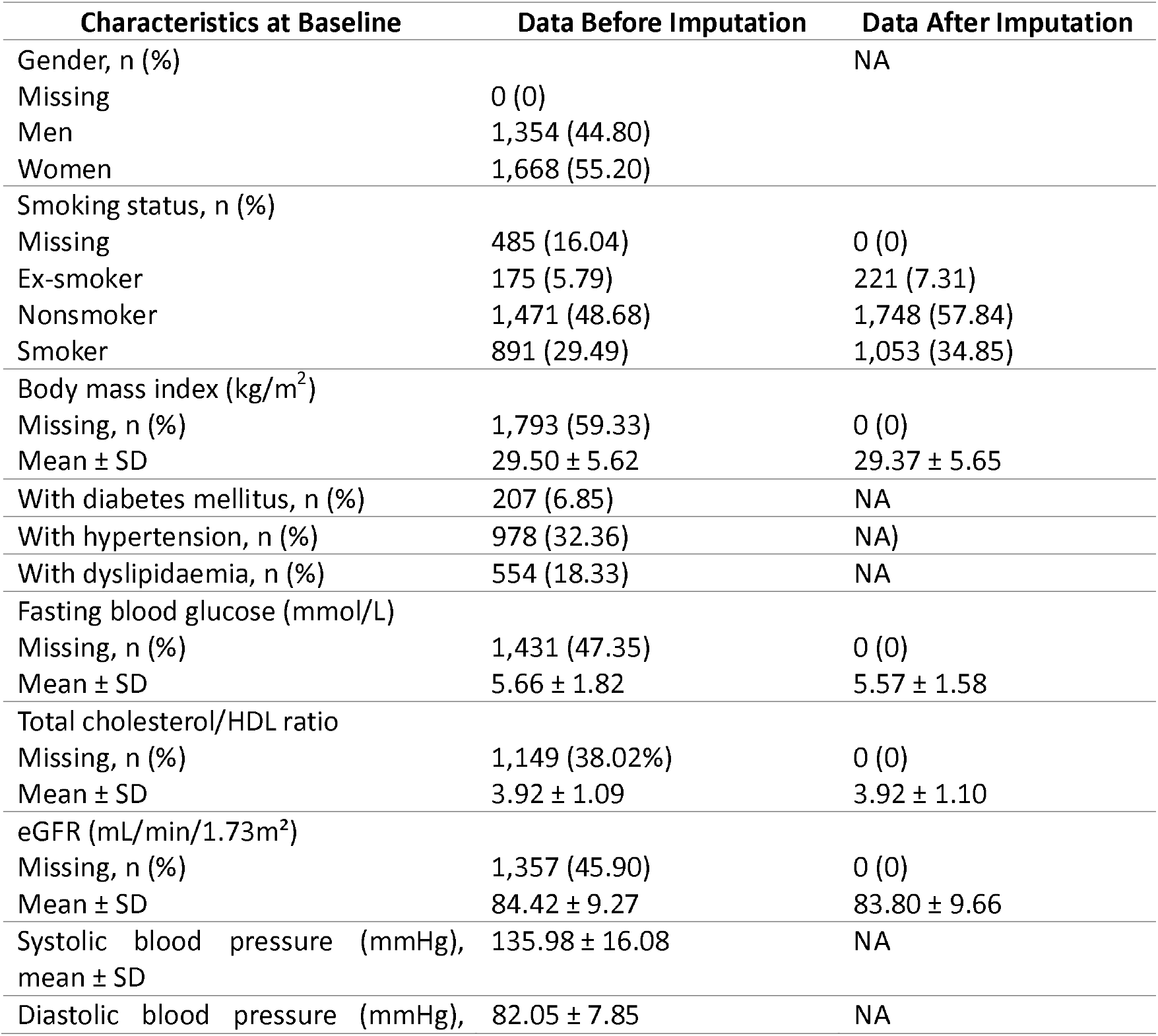
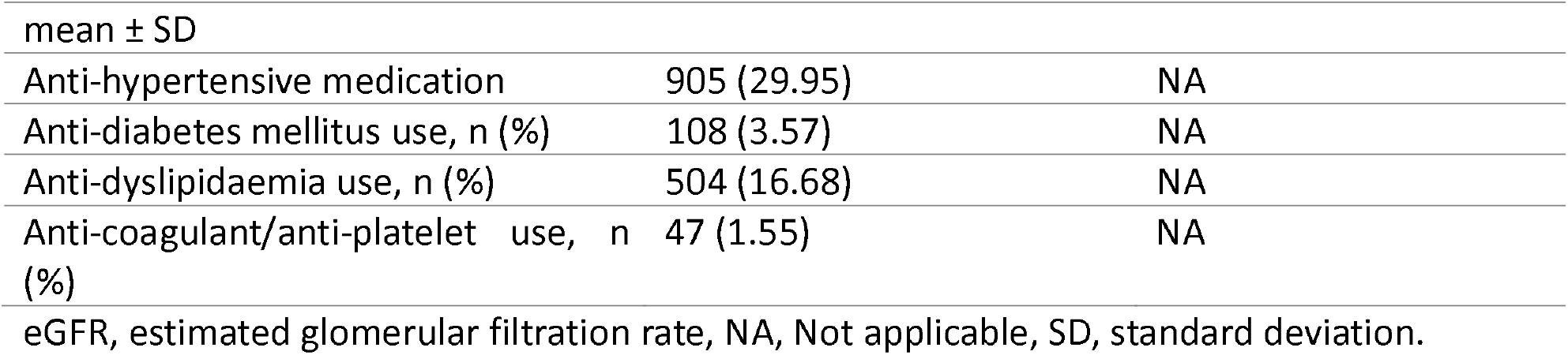
Baseline Characteristics of the Patients.

The hazard ratios of each 1-SD increase of SD, CV, and ARV are presented in Table S3. In the unadjusted model (Model 1), VVV BPV as assessed by SD, CV, and ARV was significantly associated with CVD. The SD of systolic BPV had an HR of 1.88 (95% CI: 1.71–2.08), diastolic BPV an HR of 1.94 (95% CI: 1.76–2.13). Similarly, the CV of systolic BPV and diastolic BPV were associated with HRs of 1.85 (95% CI: 1.68–2.04) and 1.96 (95% CI: 1.78–2.16), respectively. ARV indices also predicted CVD, with HRs of 1.60 (95% CI: 1.47–1.73) for systolic BPV and 1.63 (95% CI: 1.51–1.77) for diastolic BPV. Schoenfeld residuals indicated that the proportional hazards assumption was satisfied in all models (p > 0.05).

After sequential adjustment for demographic, clinical, and laboratory factors (Models 2–5), associations remained robust, with slight attenuation. In the fully adjusted model (Model 5), the SD of systolic and diastolic BPV remained associated with CVD (systolic BPV: HR = 1.71, 95% CI: 1.49–1.95, BH adjusted *p* = 2.85 × 10^−13^; diastolic BPV: HR = 1.95, 95% CI: 1.72–2.21, BH adjusted *p* = 4.35 × 10^−20^. CV measures were similarly significant (systolic BPV: HR = 1.69, 95% CI: 1.49–1.92, BH adjusted *p* = 4.18 × 10^−15^; diastolic BPV: HR =1.90, 95% CI: 1.69–2.13, BH adjusted *p* = 3.48 × 10^−21^. ARV also remained predictive of CVD risk (systolic BPV: HR = 1.36, 95% CI: 1.22–1.51; diastolic BPV: HR = 1.55, 95% CI: 1.39–1.72).

All VVV BPV metrics were significantly associated with increased CVD risk across all models, with SD consistently demonstrating slightly higher effect estimates than CV and ARV. ARV demonstrated a significantly lower HR compared to SD and CV. Adjustment using the BH procedure, which controls the false discovery rate, confirmed that these associations remained significant after accounting for multiple comparisons.

The cut-off values for SD, CV, and ARV are presented in Figure 1. The highest concordance was observed for systolic and diastolic BPV using SD, at 19 mmHg and 11 mmHg, respectively. For CV, the optimal concordance accured at 14% for systolic and 12% for diastolic BPV, while ARV showed the highest concordance at 15 mmHg (systolic) and 11 mmHg (diastolic). Detailed concordance values for all cut-offs are provided in Table S4.

**Figure 1.**
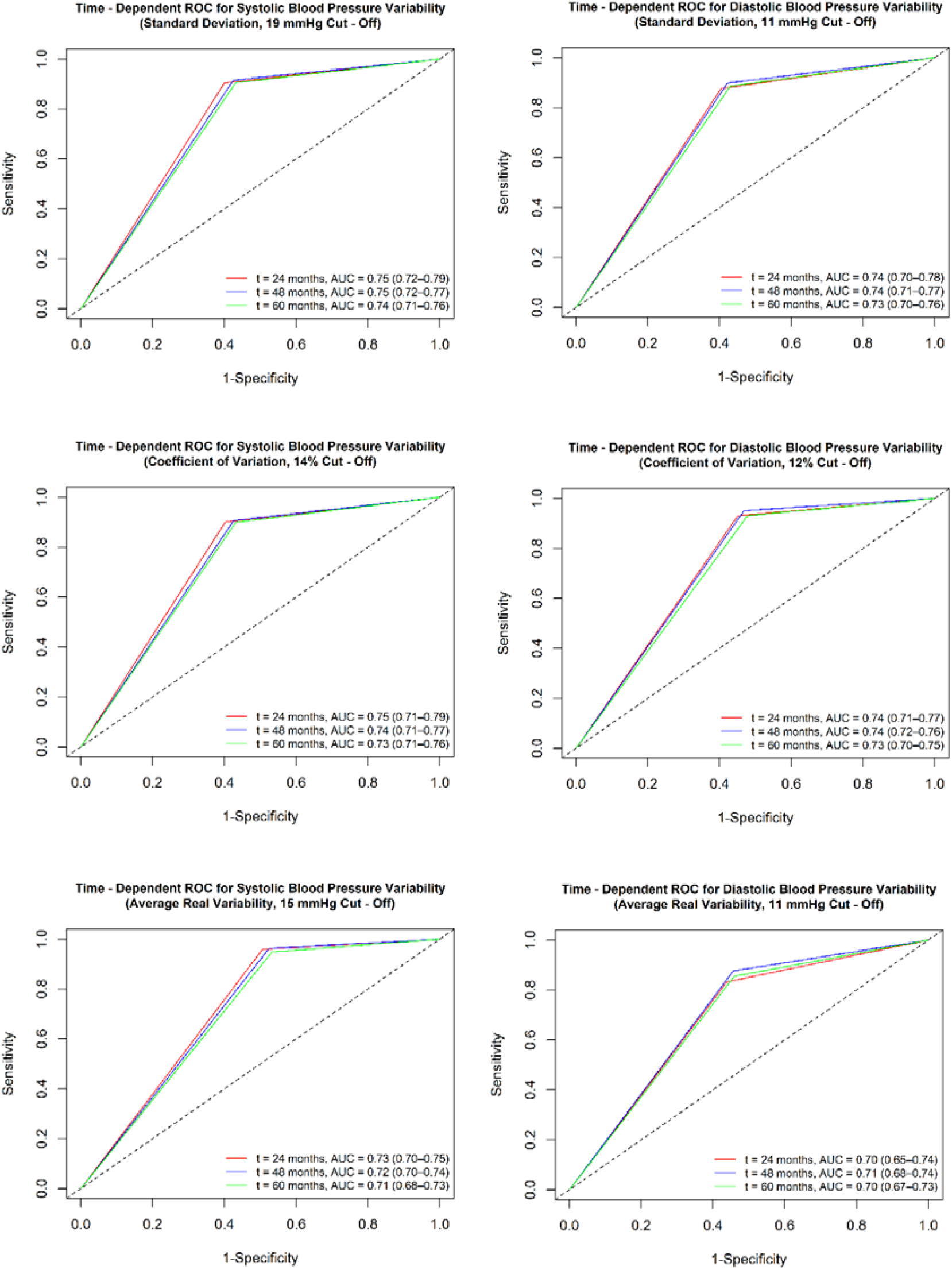
Time-dependent receiving operator characteristics of systolic and diastolic blood pressure variability for each BPV metric. AUC, area under the curve, ROC, receiving operating characteristic.

Overall, the time-varying AUC for systolic BPV was slightly higher than that for diastolic BPV across all metrics. AUC values derived from SD, CV, and ARV demonstrated similar predictive performance for cardiovascular disease over two-, four-, and five-year intervals. For the five-year period, AUC values were 0.74 (95% CI: 0.71 – 0.76) for SD, 0.73 (95% CI: 0.71 – 0.76) for CV, and 0.71 (95% CI: 0.68 – 0.73) for ARV. Diastolic BPV followed a similar pattern, with five-year AUCs of 0.73 (95% CI: 0.70 – 0.76) for SD, 0.73 (95% CI: 0.70 – 0.75) for CV, and 0.70 (95% CI: 0.67 – 0.73) for ARV.

Subgroup analysis as presented in Figure 2 revealed that patients with a mean systolic BP of <140 mmHg were at a higher risk of CVD for every 1-SD increase in BPV, as assessed by the SD BPV metric. No significant differences in HRs were observed across subgroups defined by sex, hypertension, diabetes mellitus, dyslipidaemia, or the use of antihypertensive, antidiabetic medications, or statins.

**Figure 2.**
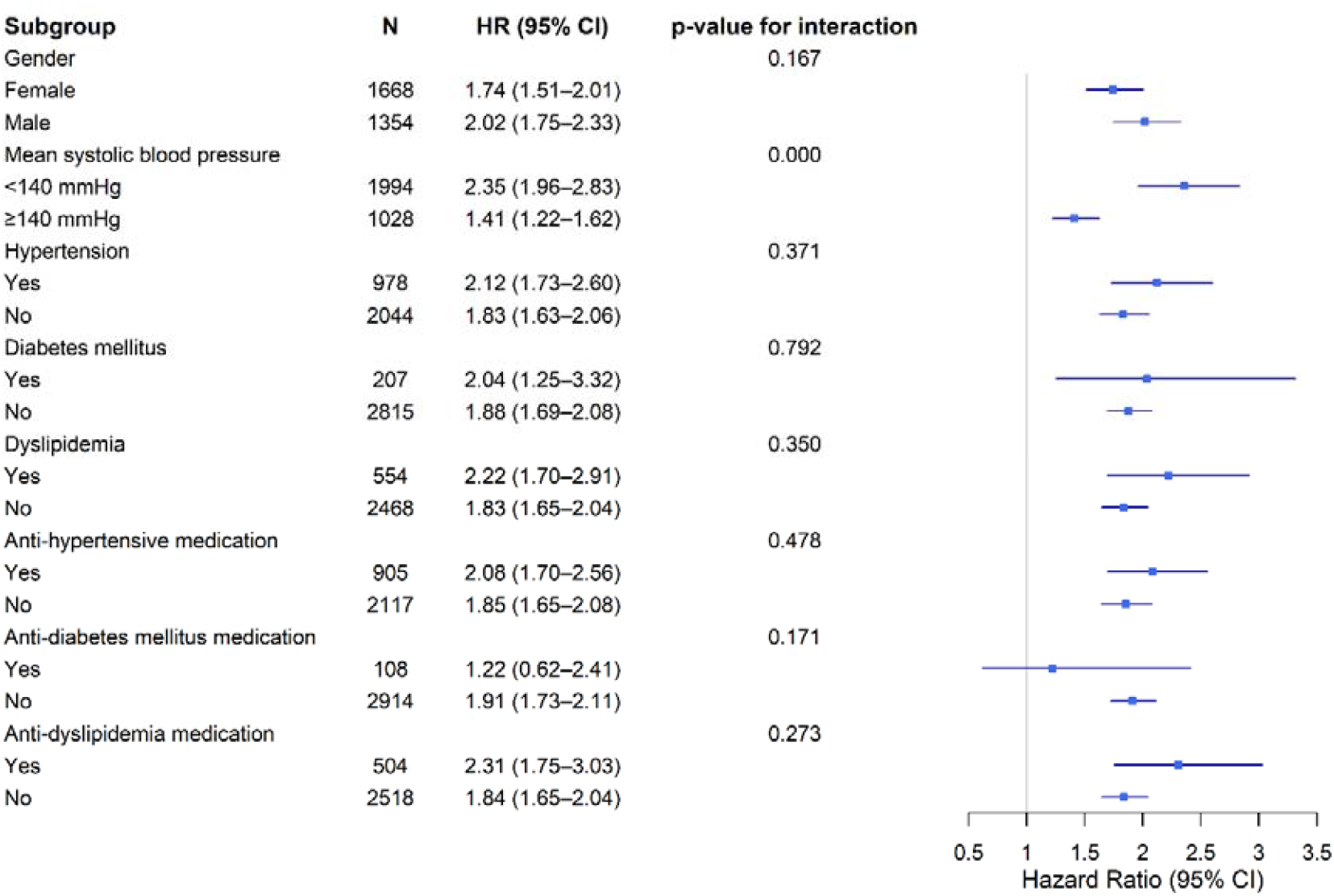
Risk of cardiovascular disease for each standard deviation increase in VVV BPV, as measured by the standard deviation, in different subgroups of patients. CI, confidence interval, HR, hazard ratio.

The Kaplan-Meier survival curves, stratified by sex and categorised by systolic BPV using a cutoff value of 19 mmHg for SD, are shown in Figure 3. The curves demonstrate distinct survival patterns based on BPV. Among participants with lower systolic BPV as measured by SD (< 19 mmHg), survival probabilities for men and women were similar throughout the follow-up period, with no statistically significant difference (log-rank p = 0.47). In contrast, among participants with higher SD (≥19 mmHg), women showed a steeper decline in survival compared with men. A clear separation between the survival curves became evident after approximately 50 months of follow-up.

**Figure 3.**
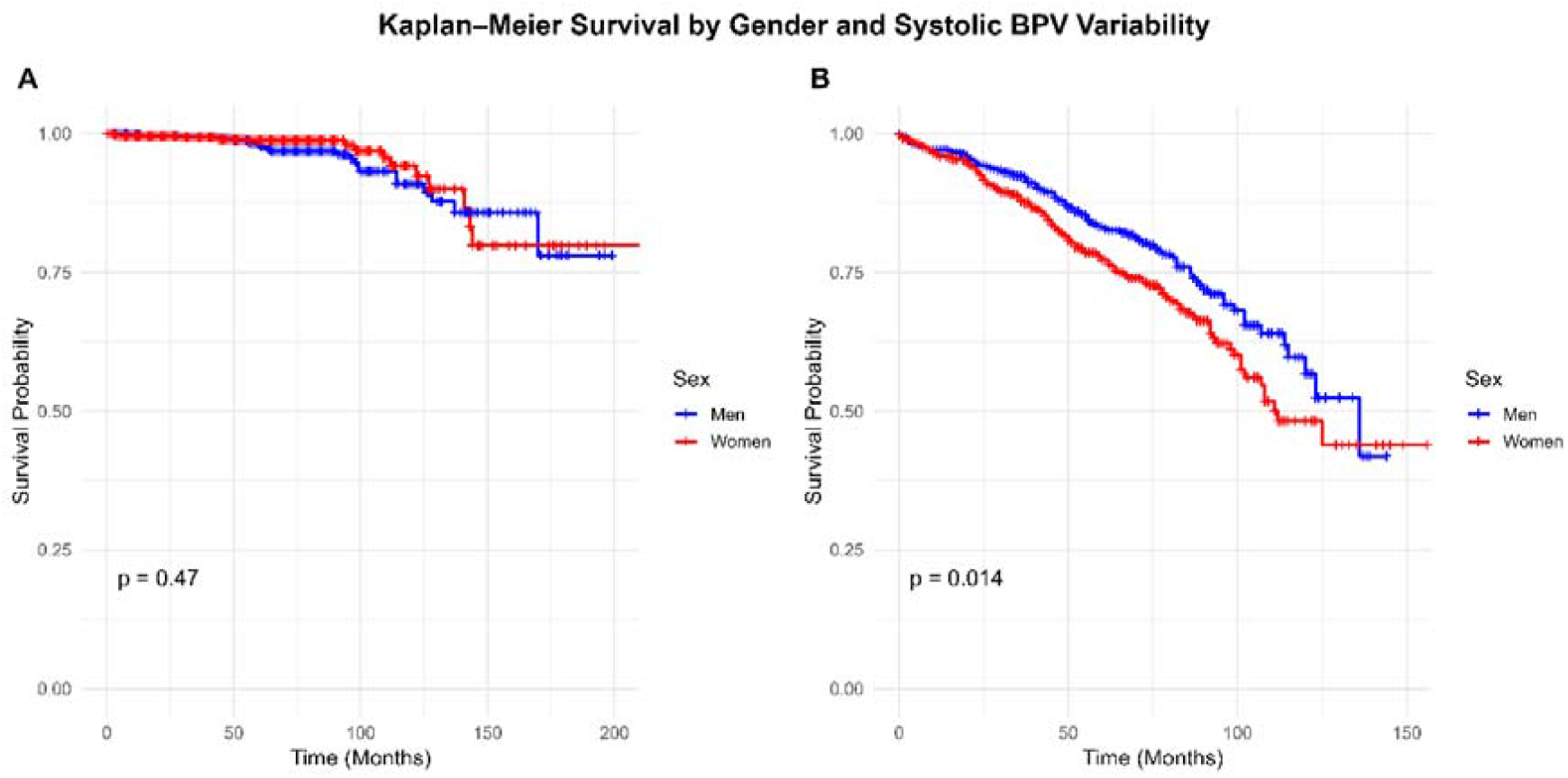
Kaplan–Meier survival curves for the first CVD event stratified by sex and the category of systolic BPV based on the cut-off value of standard deviation (19 mmHg). Figure A, standard deviation systolic BPV <19 mmHg; B, standard deviation systolic BPV ≥19 mmHg.

## Discussion

This study revealed that at least 5 BP measurements are required to calculate BPV for predicting CVD. The minimum number of BP measurements needed to estimate BPV remains a topic of debate, with limited evidence. A previous study using an GP data reported that at least six BP measurements were needed to assess BPV over a 10-year period, achieving an ICC of 0.74 when compared with 20 measurements as the reference.^14^ Obtaining even five blood pressure readings in real-world settings can be challenging, particularly among younger and mostly healthy patients. This argument is supported by a previous study that utilised MedicineInsight, a repository of EHR data from over 500 general practices in Australia, which found that only 19% of patients aged 45 to 74 had at least three visits within a two-year period.^15^ This highlights the importance of maintaining continuity of care in general practice.

In adults without a prior history of CVD, we found that VVV BPV calculated from BP measurements taken in general practice before age 55 years was positively associated with CVD events after age 55, even after adjusting for multiple confounders. To our knowledge, this is the first study to establish clinically relevant cut-off values for SD, CV, and ARV as predictors of CVD using time-varying AUC analysis over a five-year period. The time-varying AUCs for SD, CV, and ARV were approximately 0.75, indicating moderate discrimination properties. Predictive performance was slightly stronger at the two-year mark than at four or five years; however, overlapping confidence intervals and small absolute differences suggest that the identified cut-off values remain robust for up to five years. Previous evidence from a systematic review and meta-analysis of 49 studies reported a VVV BPV threshold of SD = 6.72 mmHg or CV = 9.05%, but these findings were associated with a 10% increased risk of CVD rather than actual CVD event prediction, which limits their clinical usefulness. Additionally, those studies often involved patients over the age of 60 years or those with pre-existing CVD or high-risk profiles.^16^ In contrast, our study focused on a younger, lower-risk population using real-world clinical practice data.

Previous studies have suggested that VVV BPV is an independent risk factor for CVD. A novel finding from this study indicates that individuals with a mean SBP below 140 mmHg, but high VVV BPV have a higher hazard ratio compared to those with a mean SBP above 140 mmHg. This observation has not been widely covered in the existing literature, except for an earlier study involving young African Americans. This previous study found that for individuals with a mean SBP of less than 130 mmHg and a diastolic BP of less than 80 mmHg, each 2.78-unit increase in the variability independent of the mean (VIM) of systolic BP was associated with a 25% higher risk of developing CVD (95% confidence interval: 3% - 52%).^17^ Moreover, a previous study investigating atheroma progression showed that VVV BPV was a significant predictor in the sub-cohort with a mean BP < 140/90.^18^

SD, CV, and ARV demonstrated comparable time-varying AUCs. Among these, SD remains the most used metric due to computational simplicity. Our previous meta-analysis found no significant differences in effect sizes among studies that used BPV metrics such as SD, CV, ARV, and VIM.^16^ Most previous studies have categorised BPV into percentiles, such as tertiles, quartiles, or quintiles, to predict CVD risk.^19–21^ In contrast, our study identified cut-off values for SD, CV, and ARV for both systolic and diastolic BP using time-varying AUC analysis. A previous study by Del Pinto et al defined high systolic BPV as CV > 10%, reporting a univariate HR of 1.68 (95% CI 1.28–2.21) and a multivariate HR of 1.35 (95% CI 1.02–1.80).^22^

SD and CV are established metrics for BPV that rely solely on a set of BP values, disregarding the regularity of measurements. In contrast, ARV incorporates both the sequence and regularity of the measurements, making it the optimal metric for BP data collected systematically, such as ambulatory BP monitoring (ABPM) ^23,24^. This distinction explains why ARV demonstrated a slightly lower AUC in this study, as real-world primary care practice often lacks consistent measurement intervals. Therefore, while ARV may be optimal for patients undergoing systematic BP monitoring, SD and CV are more practical and suitable metrics for estimating BPV from real-world EHR datasets.

We also found that women with higher BPV have a greater likelihood of developing CVD than men, despite having comparable risk profiles at lower BPV levels. These findings suggest that higher systolic BPV may be linked to poorer CVD outcomes in women than in men. This difference may be attributable to menopause and associated hormonal changes.^25^ Changes in estrogen levels during perimenopause and menopause have been linked to endothelial dysfunction ^26^, increased arterial stiffness ^27^, and cardiac remodelling.^28^ A landmark age of 55 years was used in our analysis, which means that many women in the female cohort may have been entering perimenopause or menopause, contributing to this difference.

### Clinical, health policy, and future research implications

The cutoff values identified in this study can be used to classify patients based on their five-year risk of developing CVD starting at age 55. Implementing a program that promotes routine BP measurement in general practice in adults, especially women aged between 45 and 55 years, coupled with public health education to raise awareness about the importance of regular BP monitoring, may identify hypertension early and provide data for VVV BPV calculation to guide early CVD prevention.^29^ Greater involvement of practice nurses and First Nations Health Workers could help improve the rate of screening for hypertension and the regularity of BP measurements. However, currently, there is no clear Medicare funding for the time required for these health professionals to perform this task. There is a health assessment item number for chronic disease screening in people aged 45 and 49 years but it can be accessed only once. The recently introduced peri/menopause item number is applicable only to women. Expanding the 45–49-year-old assessment to enable annual assessments and a wider age range could provide funding for practice nurses and the First Nations Health Workers to undertake important preventative health duties, including regular BP measurements.

This study demonstrates the value of VVV BPV can be incorporated into CVD risk score. CVD risk calculator that includes VVV BPV should be made available and ideally should be integrated into GP software. Moreover, the incorporation of VVV BPV may facilitate early stratification of CVD. A previous investigation from the CARDIA study found that VVV BPV in young adults is associated with modest adverse alteration of carotid intima-media thickness^30^, as well as myocardial structure and function, as measured by a higher left-ventricular mass index, worse diastolic function, and higher left ventricular filling pressures in later life.^31^ This suggests that VVV BPV may serve as a predictor for the development of CVD, underscoring the potential for early intervention. Further research is necessary in cohorts with lower landmark ages to substantiate the association between VVV BPV and CVD in young adults.

This study strengthens the evidence for VVV BPV in stratifying CVD risk among people with BP less than 140 mmHg, regardless of whether the patient has hypertension.^32^ VVV BPV can be applied to calculate residual CVD risk, the risk of developing CVD after achieving an optimal BP level.^33^ Therefore, it may guide pharmacological and non-pharmacological interventions for CVD prevention as suggested by previous studies.^34,35^

### Strengths and Limitations

This study utilised longitudinal BP data from EHRs in general practice, linked with hospital data, to provide valuable insights into real-world practice. The findings revealed substantial variability in the intervals between BP measurements; thus, we cannot recommend an optimal measurement interval for future CVD risk estimation. Additionally, as EHR data were used, some variables, such as risk factors, laboratory results, and body mass index, contained missing values that required multiple imputations for adjusted analyses.

We evaluated VVV BPV in each patient before age 55 to establish a clear temporal sequence between exposure and outcome, ensuring BPV assessment preceded CVD development. However, this approach may have excluded individuals who experience early-onset CVD, potentially leading to an underestimation of CVD risk in younger, high-risk individuals. Moreover, this study estimated the risk of CVD only for patients aged 55 years or older, as we had a limited number of samples from patients with a reliable number of BP records in primary care before age 55. Consequently, the proportion of smokers in this study is also higher as the consequences of having at least five blood pressure measurements in this dataset are that the patients have more chronic conditions. This fact suggested that the results of this study might not be generalisable to the general population. Further study is needed to externally validate the result.

## Conclusions

This study established threshold values for predicting CVD using systolic and diastolic VVV BPV. A minimum of five blood pressure measurements is required for calculating BPV pertinent to CVD prediction. These findings underscore the critical importance of routine BP monitoring in GP to facilitate early CVD risk stratification and prevention.

## Data availability statement

The corresponding author will make the code and data available with valid reasons and approval when required.

## Acknowledgements

We want to thank the ePBRN Primary Care Health Informatics Working Group of the Secure Research Environment for Digital Health (SREDH) Consortium (www.sredhconsortium.org, accessed on 7 October 2024) for their assistance with access to the ePBRN dataset to investigate the findings from this review. We would also like to thank Dr. Gordana Popovic and Dr. Nickson Ning from Stats Central, University of New South Wales, for their assistance during the data analysis.

## Authors’ contributions (CReDIT statement)

Conceptualization: BJ, JJ, JR, ML, RA, STL; Formal Analysis: ML; Investigation: JJ, ML; Methodology: BJ, JJ, JR, ML, RA, STL; Supervision: BJ, JJ, JR, RA, STL; Writing – original draft: ML; Writing – review & editing: BJ, JJ, JR, ML, STL; All authors read and approved the final manuscript.

## Patient consent for publication

Not applicable.

## Ethics approval

The ethics of this study has been approved by Human Research Ethics Committee of the University of New South Wales, Sydney, Australia (approval number: HC230072).

## Patient and public involvement

This study relied on secondary data from primary cares and hospitals, patients were not involved in the derivation of the cut-offs.

---

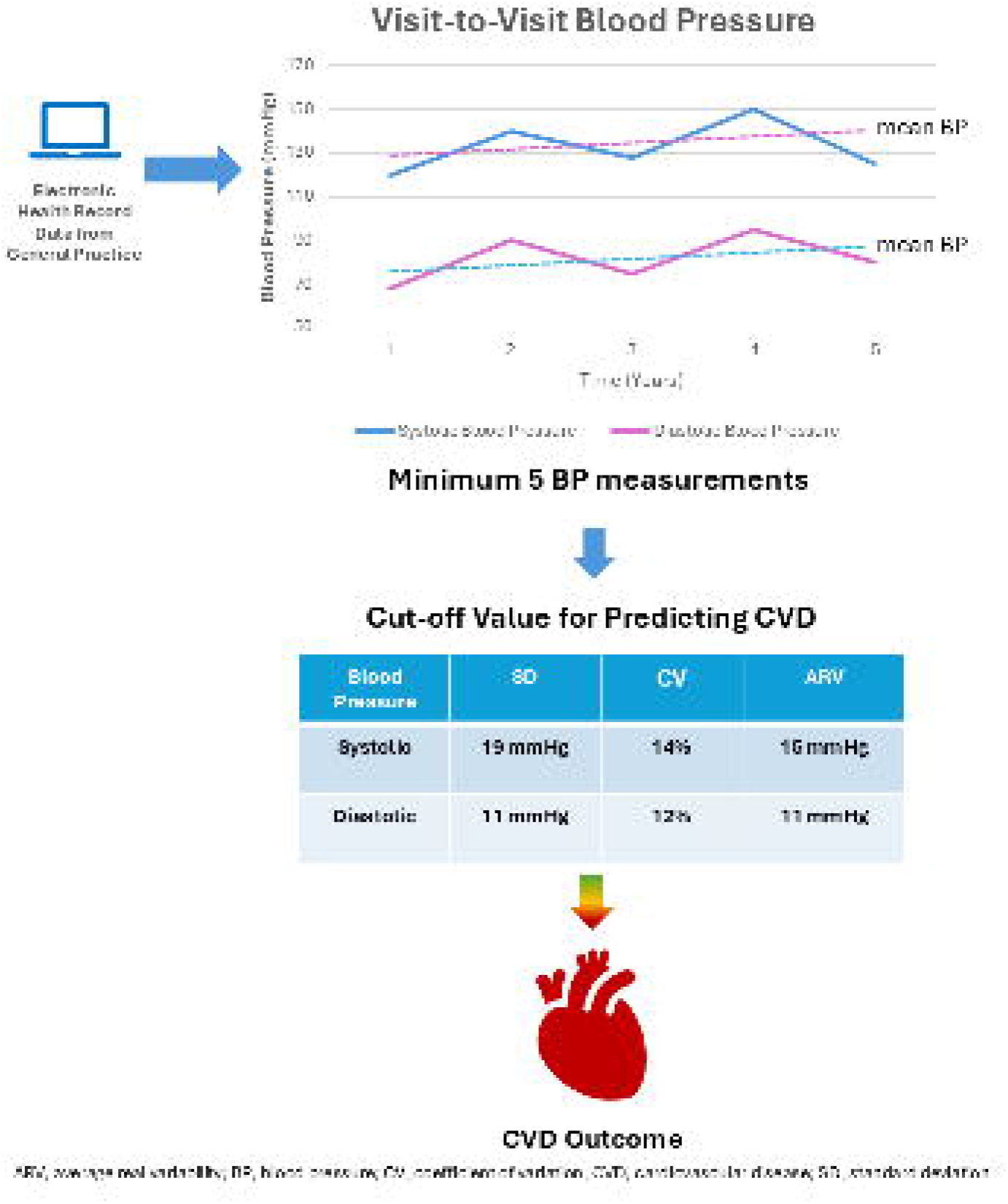

