## Supplementary Table for "The minimum number of blood pressure measurements needed and thresholds for visit-to-visit blood pressure variability to predict cardiovascular disease in primary care patients"

Supplementary methods: Details of each model developed in this study

Model 1: unadjusted

Model 2: adjusted for sex and mean BP

Model 3: adjusted for sex, mean BP, and comorbidities

Model 4: adjusted for sex, mean BP, comorbidities, and laboratory values, including total cholesterol/HDL ratio, estimated glomerular filtration rate (eGFR), fasting blood glucose, and body mass index (BMI)

Model 5: adjusted for sex, mean BP, comorbidities, laboratory values (total cholesterol/HDL ratio, eGFR, fasting blood glucose, and BMI), and medications (antihypertensive drugs, lipid-lowering agents, anticoagulants, and anti-diabetes medications).

Figure S1. The timeline of the study

**≥ 5 BP at 2 weeks apart**

**CVD event observation**

**55**

Figure S2. Flow chart of cohort selection

Assessed for eligibility

(n= 20,757)

Having at least 5 BP readings before the age of 55 years (n = 14,360)

Excluded (having fewer than 5 BP readings before the age of 55), n=6,397

Excluded (no outcome data available after the age of 55), n=10,987

Having outcome data available after 55 years old (n = 3,373)

Eligible for analysis (n = 3,022)

Excluded (having outcome before 55 years old), n=351

Excluded (having fewer than 8 BP readings before the age of 55), n=7,870

Having at least 8 BP readings before the age of 55 years (n = 12,887)

Having outcome data available after 55 years old (n = 1,900)

Excluded (no outcome data available after the age of 55), n=10,987

Excluded (having outcome before 55 years old), n=351

Eligible for analysis (n = 1,549)

= Cohort selection for the first study

= Cohort selection for the second study

Figure S3. The agreement of the standard deviation for each number of measurement (3 – 7 measurements) with 8 measurements, as assessed by intraclass correlation and Pearson’s correlation.

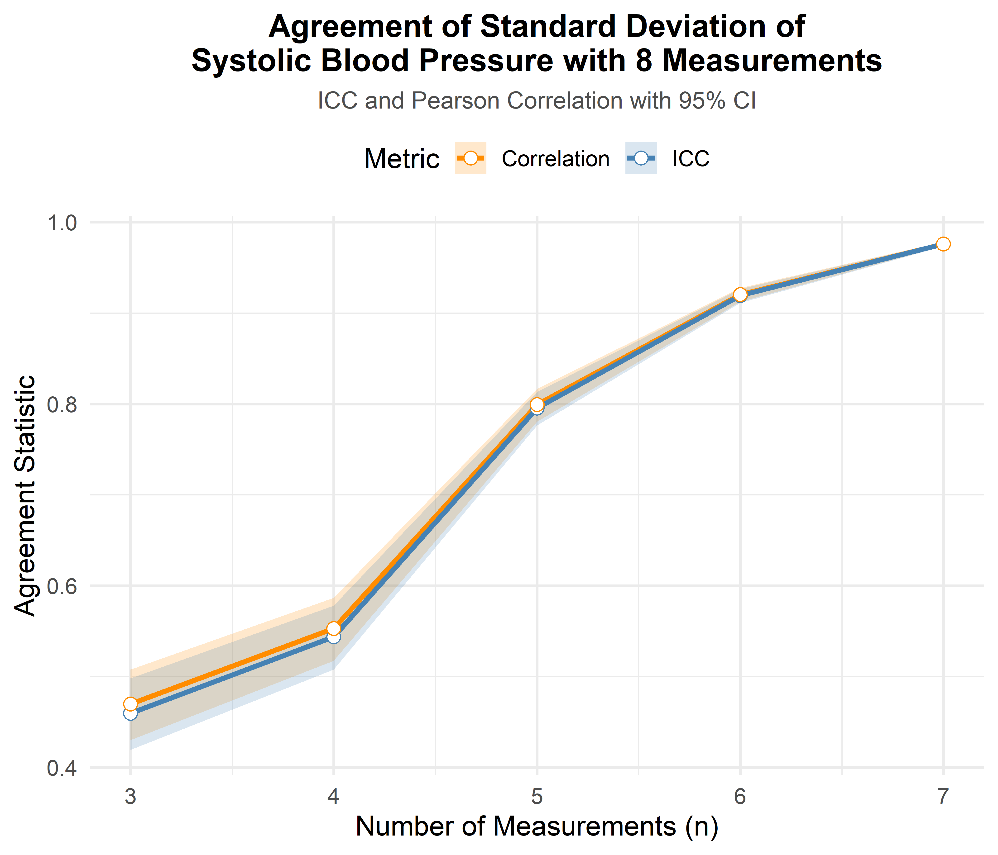

ICC values are interpreted as follows: <0.5 indicates poor reliability, 0.5-0.75 moderate reliability, 0.75-0.90 good reliability, and >0.90 excellent reliability.^9^

Table S1. TRIPOD checklist

| **Section/Topic** | **Item** | **Checklist Item** | **Page** |
| --- | --- | --- | --- |
| **Title and abstract** | | | |
| Title | 1 | Identify the study as developing and/or validating a multivariable prediction model, the target population, and the outcome to be predicted. | 1 |
| Abstract | 2 | Provide a summary of objectives, study design, setting, participants, sample size, predictors, outcome, statistical analysis, results, and conclusions. | 1 |
| **Introduction** | | | |
| Background and objectives | 3a | Explain the medical context (including whether diagnostic or prognostic) and rationale for developing or validating the multivariable prediction model, including references to existing models. | 4 |
|  | 3b | Specify the objectives, including whether the study describes the development or validation of the model or both. | 4 |
| **Methods** | | | |
| Source of data | 4a | Describe the study design or source of data (e.g., randomized trial, cohort, or registry data), separately for the development and validation data sets, if applicable. | 5 |
|  | 4b | Specify the key study dates, including start of accrual; end of accrual; and, if applicable, end of follow-up. | 5, Supplementary Figure 1 |
| Participants | 5a | Specify key elements of the study setting (e.g., primary care, secondary care, general population) including number and location of centres. | 5 |
|  | 5b | Describe eligibility criteria for participants. | 5 |
|  | 5c | Give details of treatments received, if relevant. | NA |
| Outcome | 6a | Clearly define the outcome that is predicted by the prediction model, including how and when assessed. | 5 |
|  | 6b | Report any actions to blind assessment of the outcome to be predicted. | 5 |
| Predictors | 7a | Clearly define all predictors used in developing or validating the multivariable prediction model, including how and when they were measured. | 5 |
|  | 7b | Report any actions to blind assessment of predictors for the outcome and other predictors. | 5 |
| Sample size | 8 | Explain how the study size was arrived at. | 5 |
| Missing data | 9 | Describe how missing data were handled (e.g., complete-case analysis, single imputation, multiple imputation) with details of any imputation method. | 6 |
| Statistical analysis methods | 10a | Describe how predictors were handled in the analyses. | 6 |
|  | 10b | Specify type of model, all model-building procedures (including any predictor selection), and method for internal validation. | 6 |
|  | 10d | Specify all measures used to assess model performance and, if relevant, to compare multiple models. | 6 |
| Risk groups | 11 | Provide details on how risk groups were created, if done. | NA |
| **Results** | | | |
| Participants | 13a | Describe the flow of participants through the study, including the number of participants with and without the outcome and, if applicable, a summary of the follow-up time. A diagram may be helpful. | 7, Supplementary Figure 2 |
|  | 13b | Describe the characteristics of the participants (basic demographics, clinical features, available predictors), including the number of participants with missing data for predictors and outcome. | Table 1 |
| Model development | 14a | Specify the number of participants and outcome events in each analysis. | 7 |
|  | 14b | If done, report the unadjusted association between each candidate predictor and outcome. | 7, Supplementary Table 3 |
| Model specification | 15a | Present the full prediction model to allow predictions for individuals (i.e., all regression coefficients, and model intercept or baseline survival at a given time point). | 7, Supplementary Table 3 |
|  | 15b | Explain how to the use the prediction model. | 7 |
| Model performance | 16 | Report performance measures (with CIs) for the prediction model. | 7, Supplementary Table 3, Figure 1-3 |
| **Discussion** | | | |
| Limitations | 18 | Discuss any limitations of the study (such as nonrepresentative sample, few events per predictor, missing data). | 11-12 |
| Interpretation | 19b | Give an overall interpretation of the results, considering objectives, limitations, and results from similar studies, and other relevant evidence. | 11-13 |
| Implications | 20 | Discuss the potential clinical use of the model and implications for future research. | 13 |
| **Other information** | | | |
| Supplementary information | 21 | Provide information about the availability of supplementary resources, such as study protocol, Web calculator, and data sets. | 16, Supplementary file |
| Funding | 22 | Give the source of funding and the role of the funders for the present study. | 16 |

We recommend using the TRIPOD Checklist in conjunction with the TRIPOD Explanation and Elaboration document.

Table S2. List of Medications

| **Anti-hypertensives** | **Anti-diabetics** | **Lipid lowering drugs** | **Anti-coagulants** |
| --- | --- | --- | --- |
| Natrilix SR, Dapa tabs, Lasix, Minoxidil, Natrilix, Urex M, Noten, Hydralazine hydrochloride, Norvasc, Gopten, Tritace, Cardizem, Adesan HCT, Twynsta, Exforge, Teveten lus, Diovan, Micardis plus, Micardis, Exforge HCT, Bicor, Zanidip, Lercadip, Co-diovan, Teveten, Nebilet, Avapro HCT, Coversyl, Tenormin, Coversyl plus, Atacand, Karvea, Tritace, Sevikar hct tablets, Physiotens, Sevikar, Avapro, Atacand plus, Olmetec, Norvapine, Renitec, Adalat oros, tryzan tabs, olmetec plus, vasocardol cd, cozaar, diltiazem, perindo, cordilox sr, verapamil slow release, coversyl plus ld, amlo, renitec plus, isoptin sr, sevikar hct, cardizem cd, plendil er, eplerenone, diltiazem hydrochloride, accuretic, inspra, felodur er, zestril, verapamil hydrochloride, candesan combi, tryzan, perindopril + indapamide, lasix, furosemide, betaloc, dilatrend, minax, adefin, adalat, indapamide, indapamide an, diltiazem sandoz, diltiazem an, aldactone, amiloride, prazosin, spiractin, inderal, catapres, spironolactone, verapamil, minipress, deralin, isoptin, trandate, hydrochlorothiazide amiloride hydrochloride, aldomet, pritor | Acarbose, Actos, Amaryl, Dapagliflozin, Dulaglutide, Empagliflozin, Linagliptin, Glibenclamide, Glimepiride, Glipizide, Glucagen, Hypokit, Glucagon, Glucophage, Glyxambi, Insulin, Glargine, Invokana, Janumet, Januvia, Jardiance, Liraglutide, Metformin, Nesina, Onglyza, Pioglitazone, Qtern, Rosiglitazone, Saxagliptin, Saxenda, Sitagliptin, Steglatro, Trulicity, Victoza, Apogliclazide, Apometformin, Ardix, Gliclazide, Exenatide, Isophane, Metformin | Atorvastatin, Crestor, Ezetimibe, Simvastatin, Fenofibrate, Fluvastatin, Lescol, Lipitor, Omacor, Pravachol, Pravastatin, Rosuvastatin, Vytorin, Zocor, Apo-atorvastatin, Apopravastatin, Aporosuvastatin, Aposimvastatin, Cholestyramine, Gemfibrozil, Griseostatin | Apixaban, Coumadin, Dabigatran, Etexilate, Dalteparin, Eliquis, Enoxaparin, Fragmin, Heparin, Pradaxa, Rivaroxaban, Warfarin, Xarelto, Apo-clopidogrel, Clopidogrel, Heparinoid, Prasugrel, Ticagrelor |

Table S3. Hazard ratios of each 1-SD increase of SD, CV, and ARV

| Model | BPV Metric | HR (95% CI) |
| --- | --- | --- |
| Model 1 (unadjusted) | SD of SBP | 1.88 (1.71 – 2.08) |
|  | SD of DBP | 1.94 (1.76 – 2.13) |
|  | CV of SBP | 1.85 (1.68 – 2.04) |
|  | CV of DBP | 1.96 (1.78 – 2.16) |
|  | ARV of SBP | 1.60 (1.47 – 1.73) |
|  | ARV of DBP | 1.63 (1.51 – 1.77) |
| Model 2  (Gender and mean BP) | SD of SBP | 1.67 (1.47 – 1.88) |
|  | SD of DBP | 1.96 (1.76 – 2.19) |
|  | CV of SBP | 1.65 (1.48 – 1.85) |
|  | CV of DBP | 1.91 (1.73 – 2.11) |
|  | ARV of SBP | 1.38 (1.25 – 1.52) |
|  | ARV of DBP | 1.58 (1.44 – 1.73) |
| Model 3  (Gender, mean BP, and comorbidities) | SD of SBP | 1.67 (1.47 – 1.89) |
|  | SD of DBP | 1.96 (1.75 – 2.19) |
|  | CV of SBP | 1.66 (1.48 – 1.85) |
|  | CV of DBP | 1.91 (1.72 – 2.11) |
|  | ARV of SBP | 1.37 (1.23 – 1.51) |
|  | ARV of DBP | 1.57 (1.43 – 1.72) |
| Model 4  (Gender, mean BP, and comorbidities, laboratory values, and BMI) | SD of SBP | 1.69 (1.48 – 1.93) |
|  | SD of DBP | 1.93 (1.70 – 2.18) |
|  | CV of SBP | 1.67 (1.48 – 1.88) |
|  | CV of DBP | 1.88 (1.67 – 2.10) |
|  | ARV of SBP | 1.37 (1.23 – 1.52) |
|  | ARV of DBP | 1.52 (1.37 – 1.69) |
| Model 5  (Gender, mean BP, and comorbidities, laboratory values, BMI, and medications) | SD of SBP | 1.71 (1.49 – 1.95) |
|  | SD of DBP | 1.95 (1.72 – 2.21) |
|  | CV of SBP | 1.69 (1.49 – 1.92) |
|  | CV of DBP | 1.90 (1.69 – 2.13) |
|  | ARV of SBP | 1.36 (1.22 – 1.51) |
|  | ARV of DBP | 1.55 (1.39 – 1.72) |

ARV, average real variability, BMI, body mass index, BP, blood pressure, CV, coefficient of variation, DBP, diastolic blood pressure, SD, standard deviation, SBP, systolic blood pressure

Table S4. The concordance of cut-off values for each visit-to-visit blood pressure variability metric

The concordance for each cut-off of systolic blood pressure variability

| **Standard Deviation** | | **Coefficient of Variation** | | **Average Real Variability** | |
| --- | --- | --- | --- | --- | --- |
| **Cut-off** | **Concordance** | **Cut-off** | **Concordance** | **Cut-off** | **Concordance** |
| 7.07 | 0.54 | 5.50 | 0.54 | 7.00 | 0.54 |
| 7.94 | 0.56 | 6.22 | 0.56 | 8.00 | 0.57 |
| 8.67 | 0.59 | 6.74 | 0.58 | 8.90 | 0.59 |
| 9.27 | 0.61 | 7.30 | 0.61 | 9.70 | 0.60 |
| 9.91 | 0.63 | 7.77 | 0.63 | 10.32 | 0.62 |
| 10.67 | 0.65 | 8.30 | 0.65 | 11.11 | 0.64 |
| 11.40 | 0.67 | 8.86 | 0.67 | 12.00 | 0.66 |
| 12.15 | 0.69 | 9.47 | 0.69 | 13.00 | 0.68 |
| 13.13 | 0.71 | 10.06 | 0.71 | 14.00 | 0.70 |
| 14.31 | 0.74 | 10.87 | 0.73 | 15.12 | 0.73 |
| 19.00 | 0.74 | 12.11 | 0.73 | 17.19 | 0.72 |
| 26.28 | 0.75 | 14.15 | 0.74 | 20.25 | 0.72 |
| 32.35 | 0.72 | 18.60 | 0.72 | 25.57 | 0.70 |
| 37.48 | 0.66 | 22.10 | 0.67 | 30.81 | 0.66 |
| 41.80 | 0.57 | 25.00 | 0.62 | 36.83 | 0.62 |
| 45.98 | 0.53 | 27.64 | 0.57 | 43.18 | 0.57 |

The concordance for each cut-off of diastolic blood pressure variability

| **Standard Deviation** | | **Coefficient of Variation** | | **Average Real Variability** | |
| --- | --- | --- | --- | --- | --- |
| **Cut-off** | **Concordance** | **Cut-off** | **Concordance** | **Cut-off** | **Concordance** |
| 4.58 | 0.54 | 5.80 | 0.54 | 4.29 | 0.54 |
| 5.12 | 0.56 | 6.48 | 0.56 | 5.00 | 0.56 |
| 5.59 | 0.58 | 7.02 | 0.59 | 5.62 | 0.58 |
| 6.06 | 0.61 | 7.54 | 0.61 | 6.10 | 0.61 |
| 6.49 | 0.63 | 8.06 | 0.62 | 6.62 | 0.63 |
| 6.91 | 0.65 | 8.63 | 0.65 | 7.20 | 0.65 |
| 7.40 | 0.67 | 9.22 | 0.67 | 7.69 | 0.67 |
| 7.86 | 0.69 | 9.84 | 0.69 | 8.33 | 0.69 |
| 8.39 | 0.71 | 10.59 | 0.71 | 8.90 | 0.70 |
| 9.16 | 0.73 | 11.32 | 0.73 | 9.63 | 0.70 |
| 10.04 | 0.73 | 12.37 | 0.74 | 10.83 | 0.71 |
| 11.26 | 0.74 | 14.11 | 0.71 | 12.40 | 0.70 |
| 13.47 | 0.72 | 16.51 | 0.69 | 14.32 | 0.70 |
| 16.43 | 0.69 | 19.71 | 0.64 | 17.00 | 0.69 |
| 19.01 | 0.65 | 22.34 | 0.60 | 19.86 | 0.61 |
| 21.79 | 0.58 | 25.18 | 0.59 | 23.35 | 0.57 |
| 24.37 | 0.54 | 27.96 | 0.54 | 27.49 | 0.54 |
